# Cyclical Student Shifting Models in a Limited Face-to-Face Learning Modality in the time of COVID-19 Pandemic in the Philippines

**DOI:** 10.1101/2022.08.09.22278587

**Authors:** Alexander J. Balsomo, Stephen G. Sabinay

## Abstract

**Introduction:** A joint memorandum circular (JMC No. 2021-001) was released by the Commission on Higher Education (CHED) and the Department of Health (DOH) in the Philippines, outlining the guidelines on the gradual reopening of higher education campuses for limited face-to-face classes during the COVID-19 pandemic. Besides the strict enforcement of health protocols, the said circular recommended for the reduction of COVID-19 reproduction number by limiting the number of students present within the campus through the adoption of a cyclical student shifting model. This study evaluates 32 cyclical 2-shift models and how these schedules minimizes COVID-19 incidence in the campus.

**Method:** A compartmental SEIRS model is used to simulate the number of infection for a 36-week period where the proposed 32 cyclical schedules and health protocols, such as the wearing of minimum personal protective gears and physical distancing, are inserted into the model.

**Result:** The simulation has shown that all of the proposed schedules result to low disease transmission as long as there is strong adherence to health protocols among students or strict implementation of health policies for at least 18 weeks; otherwise, the 4-10 cycle model is recommended, or shifting schedules with 3-4 consecutive weeks of in-campus classes when health protocols are poorly implemented.

**Conclusion:** The results of this study will guide university administrators in crafting their COVID-19 academic plan, especially in the selection of a suitable cyclical shifting schedule for their institution.

## 1 Introduction

During the early days of the COVID-19 pandemic in the Philippines, the Commission on Higher Education (CHED) released a series of advisories [1] to guide public and private Higher Education Institutions (HEIs) on coming up with the best decisions to manage the pandemic in their respective colleges and universities, in response to the directive of President Rodrigo R. Duterte through Proclamation No. 922 series of 2020, declaring a State of Public Health Emergency [2]. Face-to-face classes and other school-related activities were suspended, initially in the National Capital Region, and extended into whole island of Luzon, and later enforced in all parts of the country where provinces and highly-urbanized cities were given community quarantine classification [3], and the suspension of face-to-face or in-person classes were observed in areas that are at least under General Community Quarantine (GCQ).

To guide colleges and universities to implement on how to continue teaching and learning beyond the usual face-to-face instruction, CHED released a memorandum detailing the guidelines on the implementation of flexible learning [4]. Flexible learning is defined as pedagogical approach allowing flexibility of time, place, and audience including, but not solely focused on the use of technology [5]. Hence, students are to be provided with the flexibility on the learning content, schedules, access, and innovative assessment, making use of digital and non-digital tools. HEIs are autonomously given the privileges to exercise their judgement and formulate decisions to determine and implement the most viable form of flexible teaching and learning tools and strategies, determining alternative options of deliveries through various modalities, and modifying the curricular structures or program of study considering the prerequisites and corequisites of the core and elective courses. The suggested modes of deliveries are as follows: (1) online learning or blended learning, (2) macro and micro learning approach (a mix of online and offline activities), and (3) self-instructional modules; however, these modalities depend on the equipment, internet connectivity, digital literacy of instructors and students, and the learning management system and digital capabilities of the HEIs.

Although these flexible learning modalities are the safest way to deliver teaching and learning during the pandemic, improper and deficient technological and practical fundamentals have been causing problems for both teachers and students in developing countries [6]. Hence, it is expected that when COVID-19 cases will go down, eventually, teachers and students are to return to school in a cautious and gradual approach. A joint memorandum circular [7] was released by CHED and the Department of Health (DOH), outlining the guidelines on the gradual reopening of higher education campuses for limited face-to-face classes during the COVID-19 pandemic. A limited face-to-face modality refers to the on-site restriction of number of students, the implementation of a cyclical shifting schedule of classes, and the observance of health protocols such as physical distancing of at least 1.5 meters, and the proper and regular wearing of basic personal protective gears, to wit, face masks, face shields and others.

This paper focuses on the *cyclical student shifting models*. A cyclical student shifting model is a cyclic timetable where a proportion of students are allowed to be in-campus for a face-to-face instruction while others are having flexible off-campus learning modalities. CHED and DOH suggested the adoption of cyclical shifting model such as but not limited to:

1. *4-17 cycle model*. This is a 3-shift scheduling model where one-third of the students are in-campus for 4 consecutive days and out-of-campus for 17 consecutive days for flexible learning.
2. *4-10 cycle model*. This is a 2-shift scheduling model where one-half of the students are in-campus for 4 consecutive days and out-of-campus for 10 consecutive days for flexible learning.

Fig. 1 illustrates the 4-17 and 4-10 cycle models in an 18-week period. Consequently the 4-17 cycle model only allots one-third of the school calendar for a face-to-face instruction, hence, correspondingly, one-third of the instructional competencies are delivered in a regular classroom setting; while the 4-10 allots half of the school days for face-to-face delivery of half of the instructional competencies. Not all of these competencies can be effectively delivered in an online modality, for example, classes with laboratories, skills demonstration, and many others.

**Figure 1:**
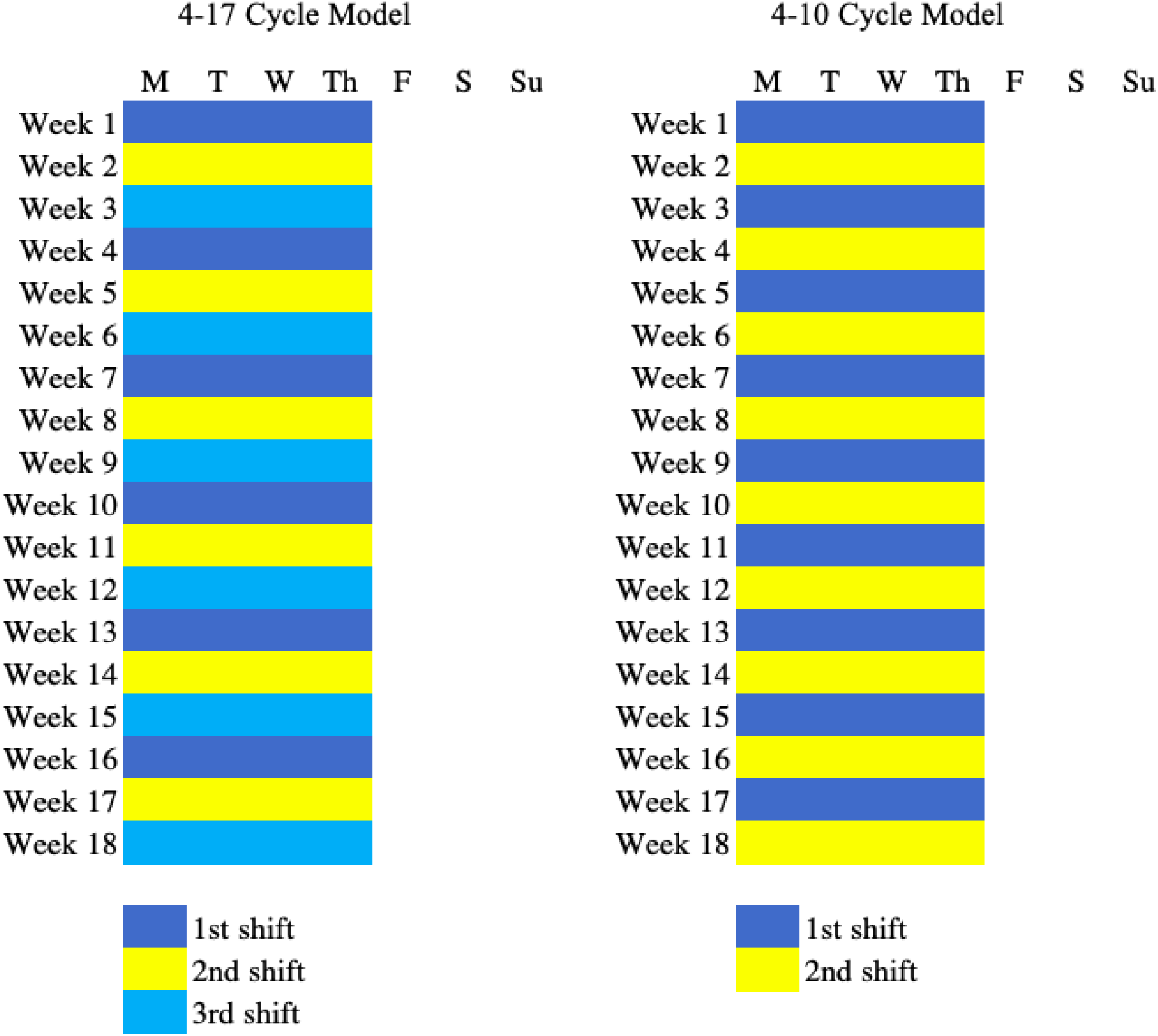
The 4-17 and 4-10 cycle models for an 18-week semester schedule

Before the pandemic, many universities in the Philippines have been adopting the standardized 5-day class block scheduling model where each course or subject is taught either on Monday-Wednesday-Friday (MWF) or Tuesday-Thursday (TTh) per week. For a 3-unit/hour lecture class, 1 hour class is scheduled every Monday, Wednesday and Friday, or 1.5 hours every Tuesday and Thursday. This scheduling has been the adopted by universities since it accommodates faculty and student preferences,and demonstrates cost advantage such as limited human and technical resources [8]. Hence, the 5-9 cycle model (5 consecutive days of face-to-face classes with 9 days of out-of-campus flexible learning) will suit these needs. But there are numerous cyclical scheduling schemes that can be explored by HEIs in order to reduce COVID-19 and/or other infectious disease cases among students.

This paper investigates different cyclical shifting models and how these models minimize COVID-19 infection within universities together with the implementation of health protocols. Our main tool is the classical compartmental Susceptible-Exposed-Infected-Recovery (SEIR) model where parameters on physical distancing and personal protection are considered. The cyclical shifting schedule, as a form of control is inserted into the compartmental model and this will be explained in the next section. Parameters on vaccination among students were not considered in this analysis due to the threat of new variants and waning of vaccine effectiveness, and likely, this exploration is applicable to many reemerging infectious disease epidemics that will likely disrupt face-to-face classes not only among the constituents of colleges and universities but as well as in the basic education sector.

Several modeling studies have been made to simulate COVID-19 infection among students in colleges and universities [9, 10, 11, 12, 13, 14] but none of these tackled the effect of cyclical shifting schedules.

This paper is organized as follows: in Section §2, the proposed 2-shift cyclical schedules are generated and the incorporation of these cyclical shifting schedules, together with level of adherence to health protocols, into the compartmental model is explained; Section §3 presents the results of the simulation of the proposed schedules with or without health protocols, and addressing the diminishing self-restraint and obedience to protocols; and finally, Section §4 summarizes the results of this work.

## 2 Methods

A compartmental susceptible-exposed-infected-recovery model is used to simulate the number of infections for a 36-week period (equivalently, two semesters without academic breaks or a 10-month basic education calendar). The schematics of the disease infection dynamics is shown in Fig. 2. The proposed cyclical shifting schedules are inserted in the model as indicator functions of the presence or absence of students in the campus.

**Figure 2:**
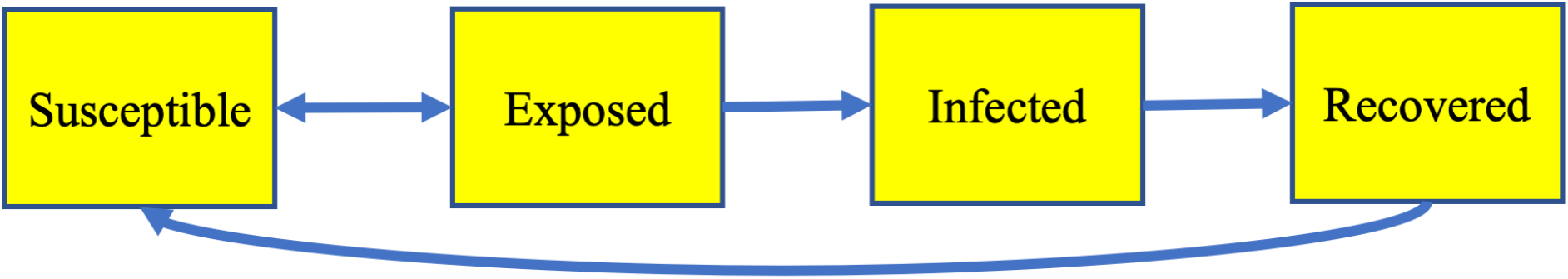
Compartmental Model

In this work, we assume that 50 percent of the school days will be attended by half of the student population in a face-to-face setting, except every Saturday and Sunday. Hence, all the proposed cyclical shifting schedule are 2-shift schedules and the simulation will only be applied to students in the 1*st* shift. In this case, we suppose that there are 10 thousand student population (*N* = 10, 000) in each shift.

### 2.1 The model

The compartmental model, shown in Fig. 2, is described by the following system of differential equations:

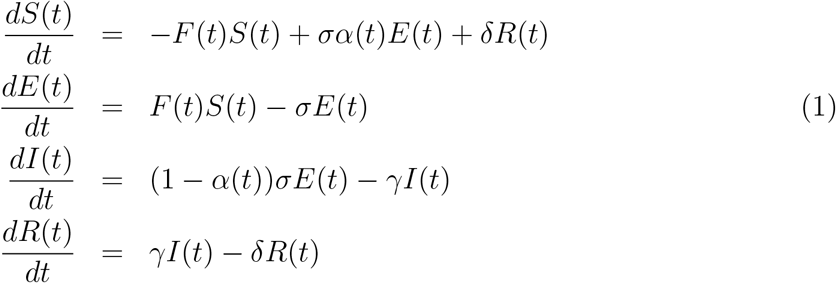

where *S, E, I* and *R* are the number of susceptibles (those who are susceptible to the virus), exposed (those who are exposed to the virus but not capable of transmitting the virus), infected (assumed to be capable of transmitting the virus to the susceptibles), and recovered individuals or those who have recovered from the disease. This model extends the *SEI* model proposed in [15] into *SEIRS* where COVID-19 reinfection after 90 days is possible [16] as determined by the rate of reinfection *δ*. The parameter *α*(*t*) is the rate of an exposed individual becoming not infected through personal protective equipment (PPE) such as proper wearing of masks and other protective gears. This is a function of time *t* since student personal protection practices can vary throughout the school period.

In [17], the incidence rate is described by an exponential decay function on the number of infectious cases. In a similar manner where distancing is considered, the frequencydependent force of infection *F* is given by

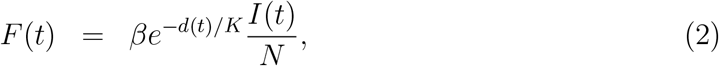

where *d* is the physical distance (in meters) and *K* is the maximum distance that can be possibly implemented in the campus. This practice of staying away from each other has been one of the health protocols mandated to the public at the beginning of the pandemic. Since *d/K* can be thought of as the proportion of the susceptible population protected from the virus through physical distancing measures, the product *βe*^−*d*(*t*)*/K*^ of transmission *β* and contact probabilities measures the rate of transmission of the virus from infected individuals to the susceptible population. Hence, *α*(*t*) and *d*(*t*) are the parameters describing health protocol which are functions of time while the parameters *β, σ* and *γ* are, respectively, the usual transmission, incubation and recovery rates.

Since the model assumes that the infected cases among students are asymptomatic [18, 19, 20, 21] and undetected [22], hence, there are no isolation for infected and exposed students. We assume that the student population is fixed with no deaths.

### 2.2 Cyclical shifting schedules

To generate the proposed 2-shift cyclical schedules for a limited face-to-face instruction, let us fix that students in the 1*st* shift will be in-campus every Monday at the beginning of each cycle, and each cycle is either 180°-rotational invariant or horizontal reflection invariant regardless of the shifts, and no in-campus or off-campus flexible learning activities are conducted every Saturday and Sunday. This idea is illustrated in Fig. 3 where the 1*st* shift is shaded blue.

**Figure 3:**
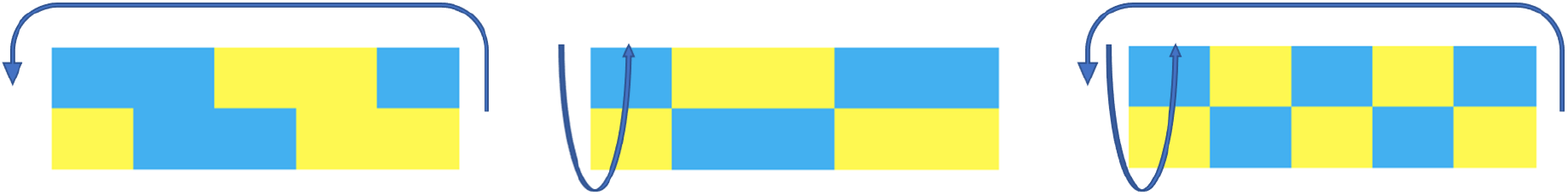
3/2-day cycle schedules that have 180°-rotational invarant (left), horizontal reflection invariant (middle), and both (right)

The proposed schedules are classified as follows:

1. *m/n*-day cycle. There are *m* days of in-campus face-to-face classes and *n* days asynchronous classes every 1*st* week in a 2-week cycle. Hence, we have the following schedules: 1*/*4-day cycle, 2*/*3-day cycle, 3*/*2-day cycle, 4*/*1-day cycle, and 5*/*0-day cycle (or 5-9 cycle model).
2. 2-week cycle. In-campus face-to-face classes are conducted in 2 consecutive weeks except on weekends and to be followed by 2 consecutive weeks of out-of-campus asynchronous activities.
3. 3-week cycle. In-campus face-to-face classes are conducted in 3 consecutive weeks except on weekends and to be followed by 3 consecutive weeks of out-of-campus asynchronous activities.
4. 4-week cycle. In-campus face-to-face classes are conducted in 4 consecutive weeks except on weekends and to be followed by 4 consecutive weeks of out-of-campus asynchronous activities.

There are 31 cyclical shifting schedules that have been generated: 2 schedules with 1/4day cycle, 7 schedules with 2/3-day cycle, 11 schedules with 3/2-day cycles, 7 schedules with 4/1-day cycle, 1 schedule with 5/0-day cycle, and 1 schedule for each 2-week, 3-week and 4-week cycles (see the Appendix).

Each of these cyclical shifting schedules is modelled by the indicator function

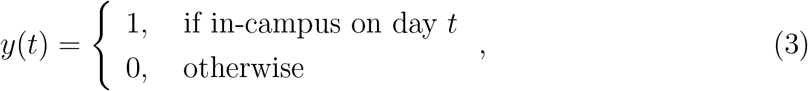

inserted into the force of infection *F* in (2). The compartmental model in (1) is now given by

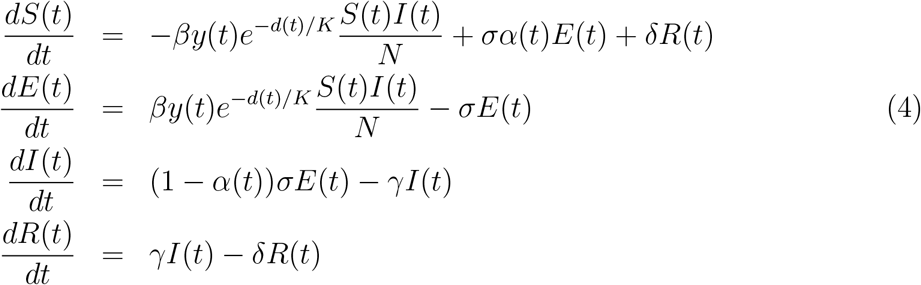

### 2.3 Personal protection and physical distancing

In [15], health protection via PPE is assumed to remove a substantial fraction of the risk faced by frontliners in hospital setting, regardless of the number of COVID-19 patients, number of encounters and exposure time. These measures are a combination of face masks, eye protection, biohazard suits and others. In this paper, we define the protection level *α*(*t*) as the fraction of the infection risk being removed or mitigated by students via PPE within the campus at time *t*; hence, *α*(*t*) ranges from 0 to 1.

On the other hand, physical distancing *d*(*t*) ranges from 0 to *K* where a maximum physical distance *K* depends on the physical features of the campus and the density of the student population.

Although students mostly adhere to COVID-19 guidelines [23], however, self-restraint in observing these protocols is expected to decrease over time [24]. In this study we assume that *α*(*t*) and *d*(*t*) are non-increasing functions, in particular, obedience and adherence to these protocols are modelled by the logistic decay functions

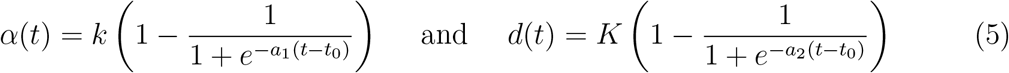

where *k* and *K* are the maximum personal protection level and physical distancing, respectively; *a*_1_, *a*_2_ are the intrinsic decay rates, and the point of inflection on the logistic curve is at *t*_0_ days. The logistic decay functions assume that self-restraint and obedience to the protocols are sustained up to *t*_0_ days, but these behaviors exponentially drop when these guidelines are poorly maintained.

Fig. 4 shows the simulation of *α*(*t*) with a protection level of at most 50% at the beginning of the school days but adherence to wearing PPE are weak after 30 days while *d*(*t*) simulates students’ physical distancing of at most 3 meters but neglects the said mandate after 126 days since its implementation at the start of the semester. This paper assumes that *t*_0_ is the same for both *α*(*t*) and *d*(*t*)

**Figure 4:**
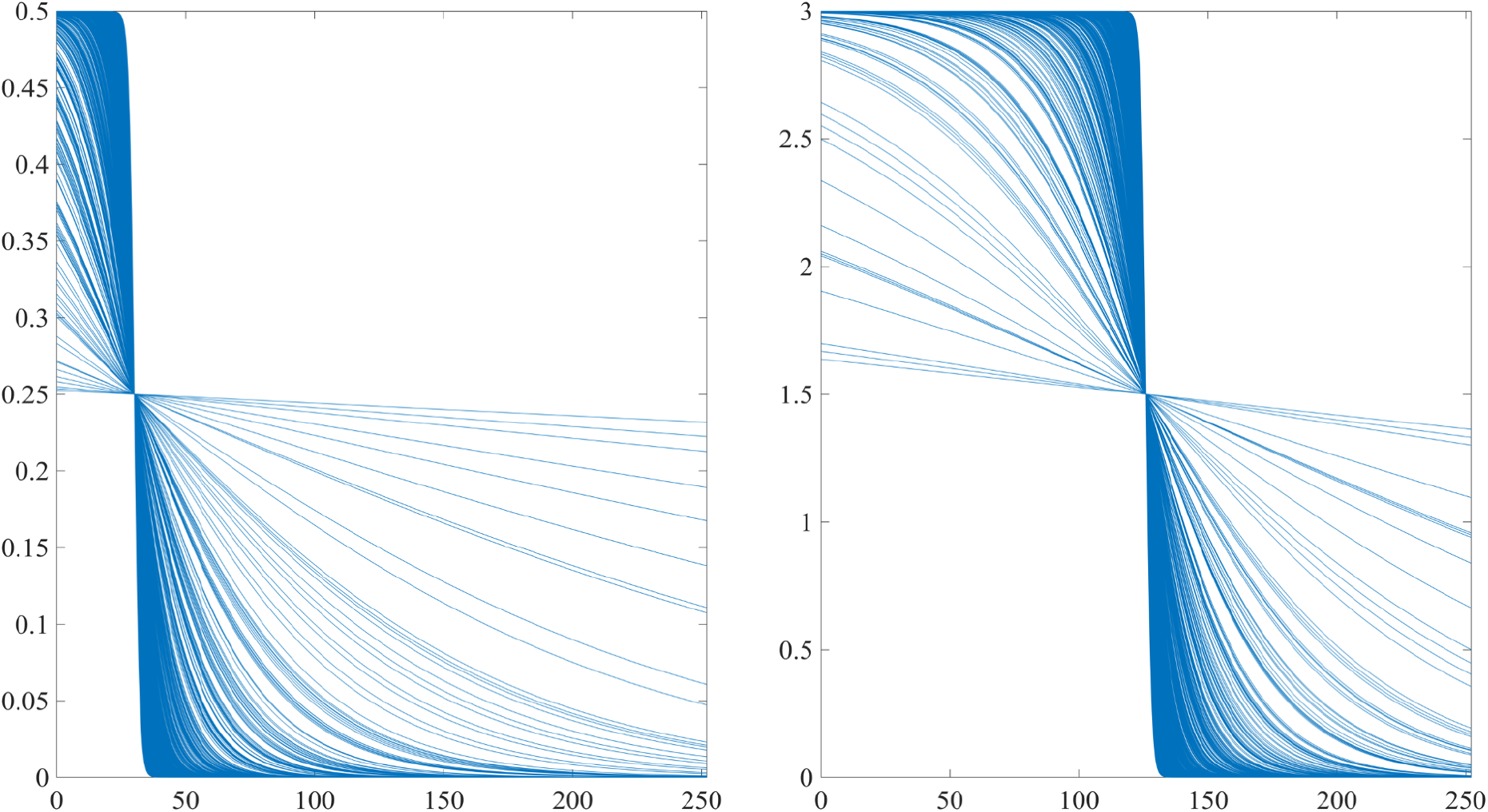
Simulated *α*(*t*) with *k* = 0.5 and *t*_0_ = 30 days (left), and *d*(*t*) with *K* = 3 meters and *t*_0_ = 126 days (right) where *n* = 1000 with *a*_1_, *a*_2_ uniformly distributed on [0, 1]

### Simulation

For each cyclical student schedule, model (4) is simulated *n* = 1, 000 times over the following uniformly distributed COVID-19 parameters (basic repreduction number between 2 and 4 [25], incubation period between 4 and 7 days [26], recovery period between 14 and 24 days [27], and COVID-19 reinfection after 90 days but not later than 300 days [28]), and health protection functions such as *α*(*t*) and *d*(*t*). The simulation using Euler’s method with a step size of 1/48 or 0.5 hours per day will be implemented using MATLAB (version R2021b) in a desktop computer (Windows 10 OS with Intel Core i7 and 16GB DDR4 3200). The link to the MATLAB code is found in the Supplementary Materials. For the initial conditions of model (4), suppose that the student population is *N* = 0, 000 with *I*_0_ = 10 asymptomatic COVID-19 infected individuals. The number of initial exposed individuals to an infected contact is Poisson distributed with mean 6.181978, taken from the contact matrix for ages 16-20 years old in the Philippines [29].

We evaluate the proposed scheduling policies including the 4-10 cycle model by calculating the expected number of total infected students at the end of the period for each cyclical student schedule. We also look at the linear relationship of the active cases via the Pearson Product Moment Correlation *R* of the proposed schedules with 4-10 cycle model. Since the daily incidence is determined, the average daily cases and the 7-day incidence rates (IRs) per 100,000 are also calculated and illustrated. Since no hospitalization nor isolation were assumed, these IRs are evaluated with the old CDC framework on COVID-19 community level indicators: low transmission if 0-9 cases, moderate transmission if 10-49 cases, substantial transmission if 50-99 cases and high transmission if *≥* 100 cases [30].

## 3. Results and Discussion

### 3.1 Simulating schedules with no health protocols

Suppose *α* = 0% and *d* = 0 meters, i.e. no health protocols are to be implemented in-campus. Fig. 5 shows that the 4-10 cyclical schedule would be an obvious choice to reduce COVID-19 incidence in the absence or less effective implementation of COVID-19 health protocols.

**Figure 5:**
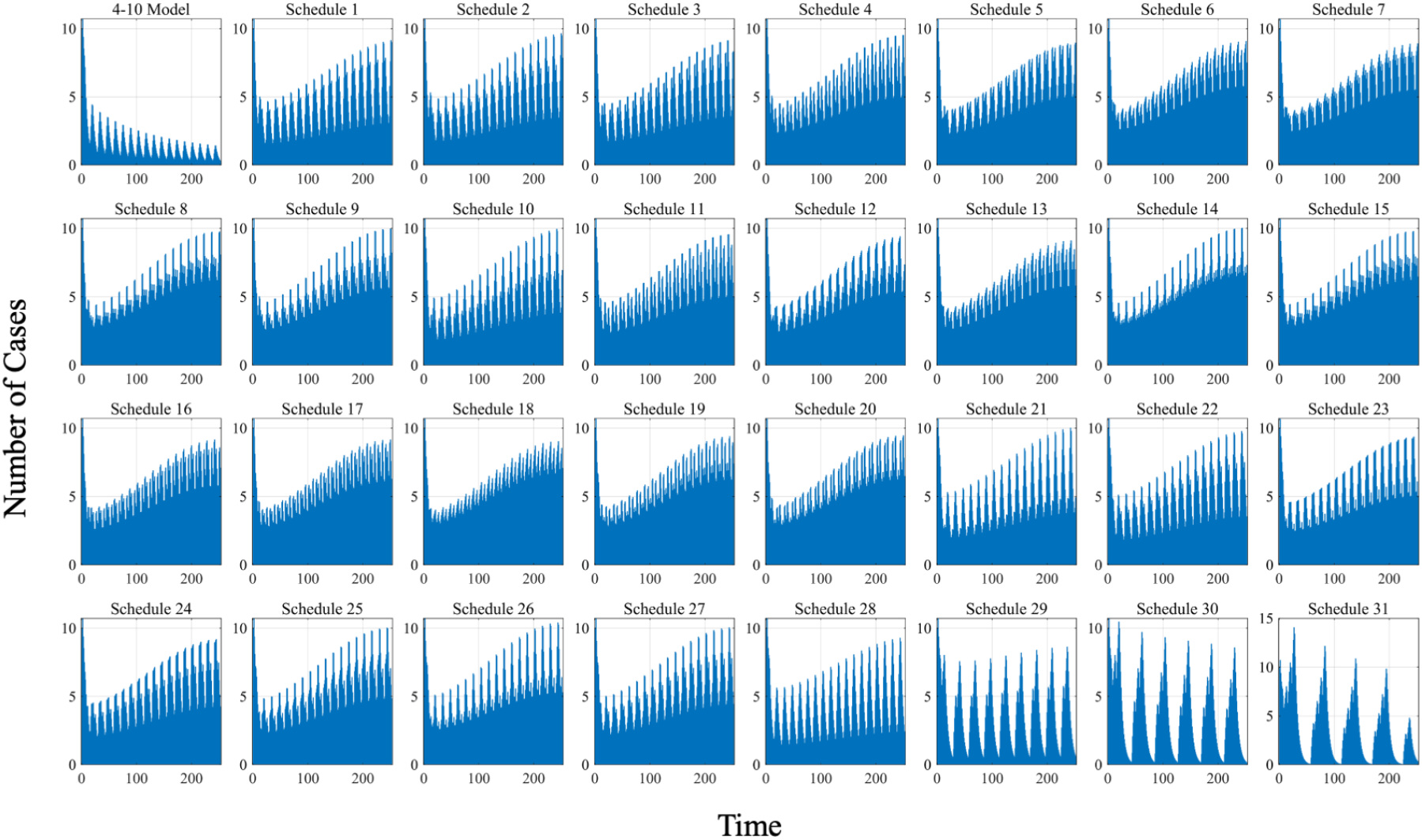
Average daily COVID-19 incidence for each cyclical schedule where no health protocols are implemented in-campus (*α* = 0% and *d* = 0 meters)

This is indeed an appropriate suggestion for a cyclical student shifting model, supporting the joint CHED-DOH policy [7]. However, the 4-10 cycle model is a 4-day block schedule and since many universities in the Philippines have been adopting a 5-day time block class scheduling before the pandemic, it is deemed appropriate to investigate other cyclic student shifting models such as the proposed *m/n*-day cycle models (Schedules 1-28), and the 2-week (Schedule 29), 3-week (Schedule 30) and 4-week (Schedule 31) cycle models- these models allot 50% of the school days for face-to-face synchronous instruction. In the same figure, we can see that the 3-week cycle and 4-week cycle models show decreasing trend of daily COVID-19 incidence in less frequent but high surge of cases compared to the 4-10 cycle model. However, the cumulative number of cases at the end of 36 weeks (or 252 days) is 402 for the 4-10 cycle while 945 and 981 cases for the 3-week cycle and 4-week cycles models, respectively.

In Fig. 6, all the *m/n*-day cycle (Schedule 1-28 *R < −*0.75) and the 2-week cycle (Schedule 29 *R* = *−*0.23) models negatively correlate with the 4-10 cycle model. Since there is a decreasing trend of COVID-19 incidence rates with the 4-10 cycle model despite the absence of health protocols, COVID-19 incidence rates with respect to Schedules 1-29 are increasing. Although the 3-week cycle (Schedule 30 *R* = 0.26) and 4-week cycle (Schedule 31 *R* = 0.46) positively correlate with the 4-10 cycle model, the surge of cases fluctuates with much higher amplitudes in these models compared to the 4-10 cycle model. Moreover, except for the 4-10 model which lead to a substantial disease transmission (*IR >* 50), all of the proposed cyclical schedules will lead to more often high disease transmission (*IR ≥* 100) throughout the school year. At the end of 36 weeks (or 252 days), there are at least 1,110 cases with *m/n*-day cycle models.

**Figure 6:**
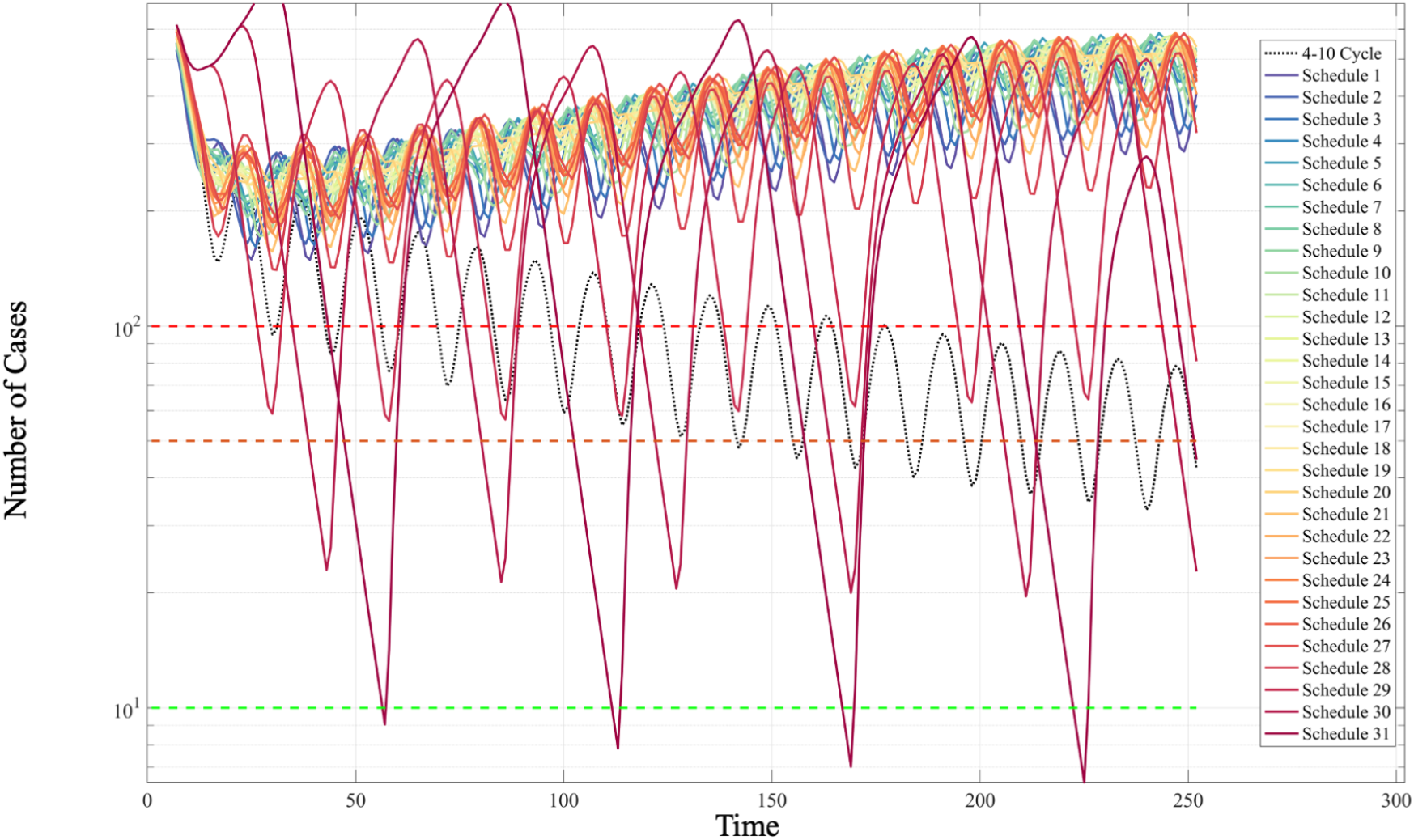
7-day IRs per 100 thousand for each cyclical schedule where no health protocols are implemented in-campus (*α* = 0% and *d* = 0 meters)

These results corroborate the work in [31] on timing a complete lockdown days before an epidemic to peak where significant decrease is appreciable if lockdowns are declared closely before the peak of a surge. Our simulation, especially for Schedules 30-31 where asynchronous activities are implemented every 3-4 weeks and evidence of COVID-19 surge, has shown decreasing trends if students are allowed to have longer days on having face-to-face instruction, and consequently longer off-campus alternative instruction. This has been a risky alternative since infection can still take place outside school premises if students consider these asynchronous off-campus activities as long academic breaks and return to campus as infecteds [14]. This paper assumes that all students stay in a safe environment with very limited human interaction every time they are outside the campus. Hence, the 3-week and 4-week cycle models are better alternatives to the 4-10 cycle model when health protocols are poorly implemented or when many students will not heed public health warnings within the school premises.

### 3.2 Simulating schedules with health protocols

Now we assume that personal protection is at *α* = 20% while physical distancing is the recommended *d* = 1.5 meters; and these protocols are strictly implemented for 36 weeks or 252 days. As shown in Fig. 7, any of the proposed cyclical schedules is recommended. The combination of these non-pharmaceutical interventions (NPIs) has led to a significant decrease of cases where there are only 100-150 simulated cases for all the schedules at the end of the school year. At the middle of the school year, low disease transmission (*IR <* 10) is observed for all models, as shown in Fig. 8, although the disease incidence rates of the 4-10 cycle model decreases faster than incidence rates of Schedules 1-28 or the *m/n*-day cycle models. But all these proposed schedules have strong positive correlation with the 4-10 cycle model (*R >* 0.95).

**Figure 7:**
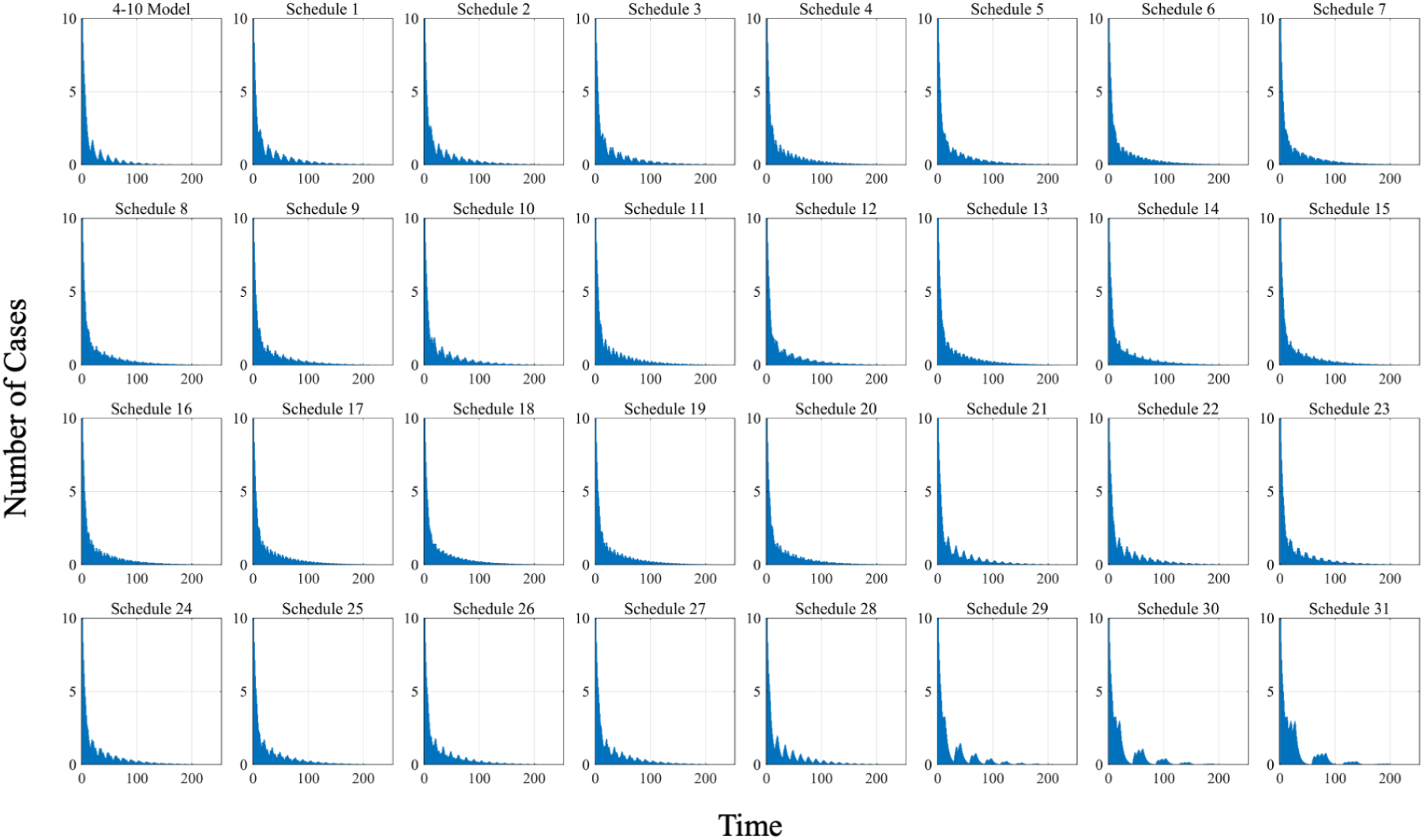
Average daily COVID-19 incidence for each cyclical schedule where *α* = 20% and *d* = 1.5 meters

**Figure 8:**
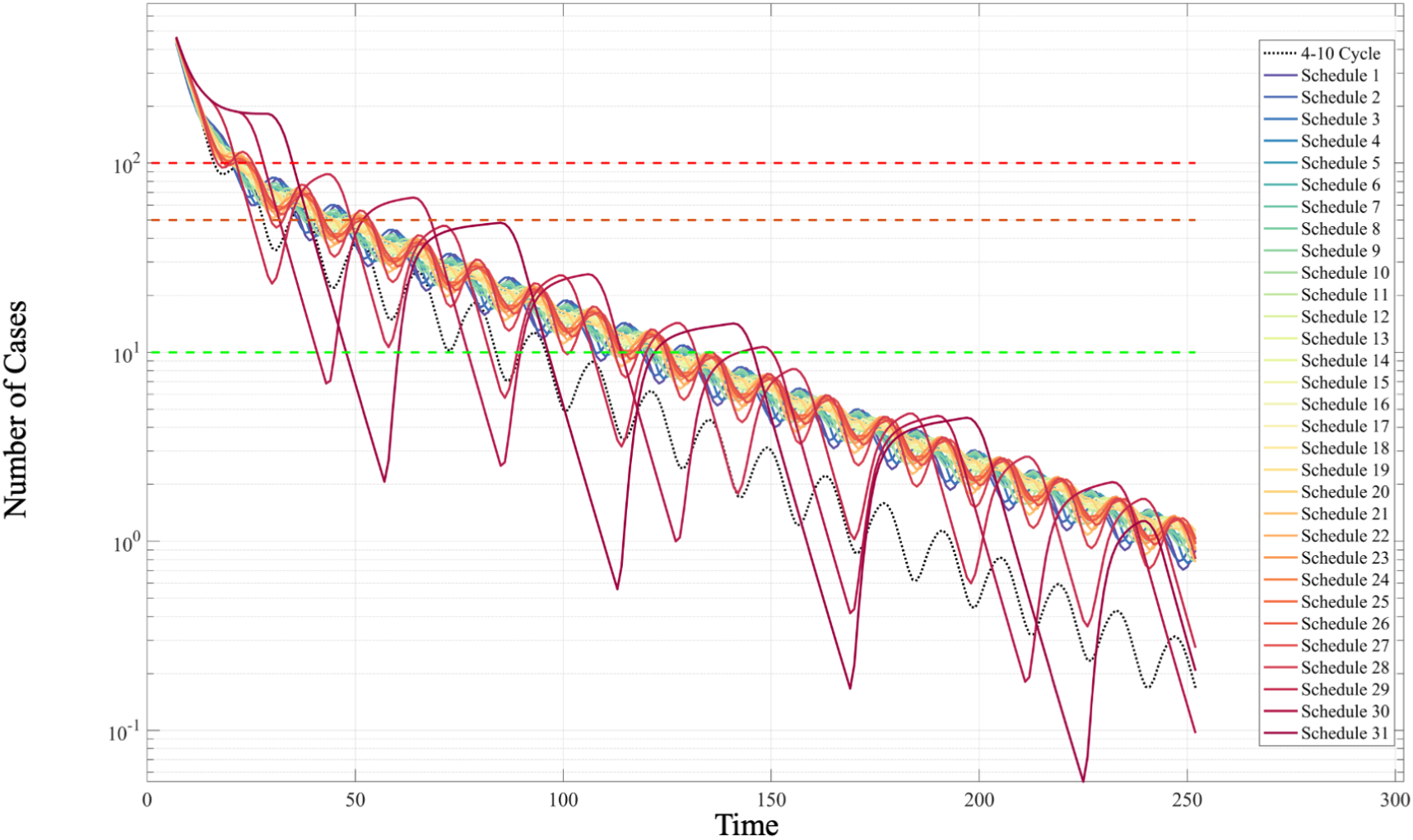
7-day IRs per 100 thousand for each cyclical schedule where *α* = 20% and *d* = 1.5 meters

If personal protection *α* ranges from 0% to 40%, or equivalently, from wearing no mask to wearing N95 masks, while physical distancing ranges from 0 to 2 meters, then the combination of interventions and the level of implementation can provide immediate impact on COVID-19 incidence. This is shown in Fig. 9.

**Figure 9:**
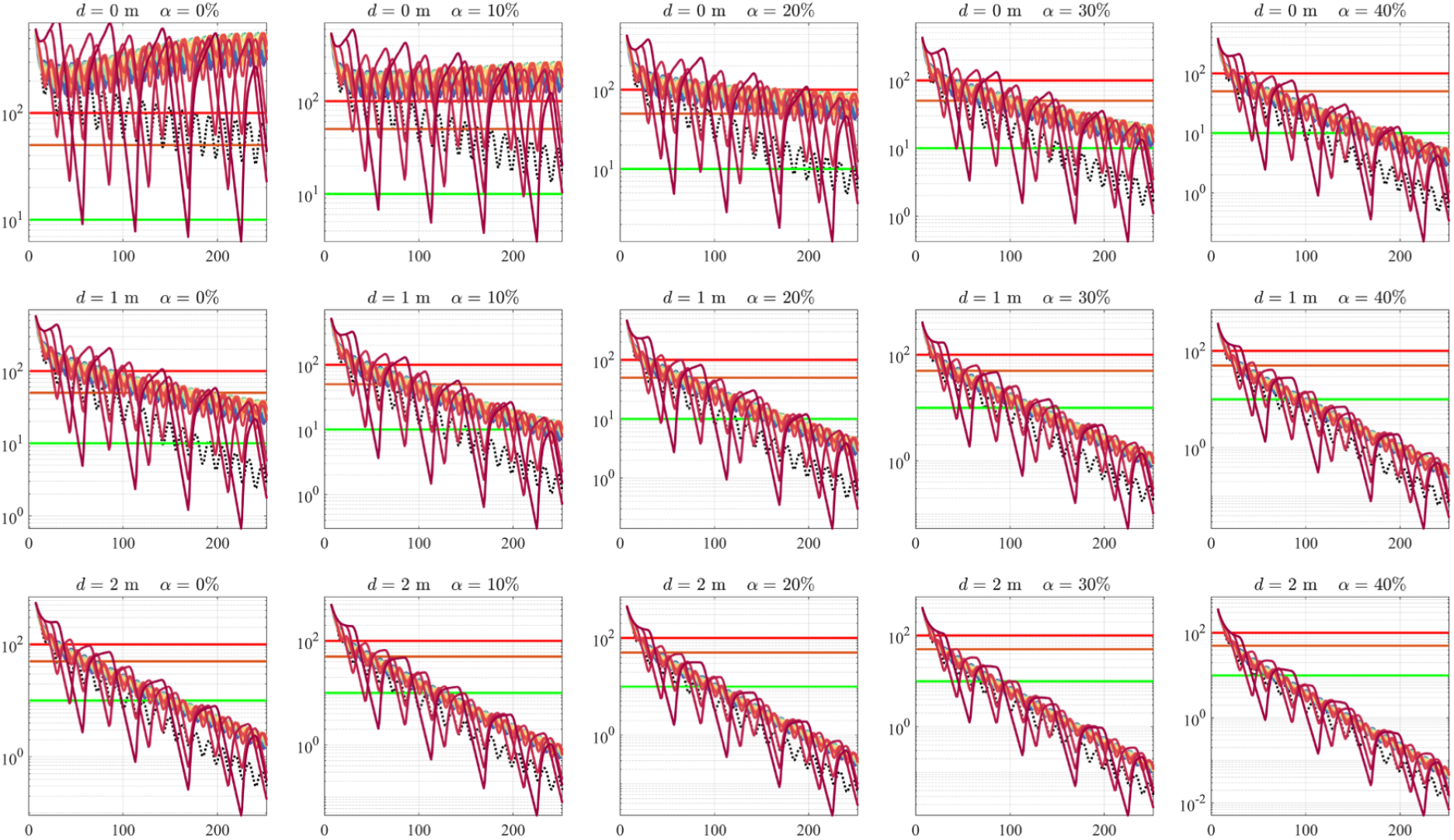
7-day IRs per 100 thousand for each cyclical schedule where *α* = 0% to 40% and *d* = 0 to 2 meters

NPIs as control measures are known to effectively reduce COVID-19 when compliance is high [32, 33, 34]. These measures has been adapted in the Philippines as minimum health standards under the DOH’s “BIDA Solusyon sa COVID-19” campaign [35] where the ‘B’ and ‘D’ in the acronym refer to mask mandate and 1-meter physical distancing, respectively, and it continues to be enforced by the Philippine government as of this writing until the State of Public Health Emergency will be lifted [2].

### 3.3 Simulating schedules with student behavior

Despite the belief that adherence to health protocols is not difficult to perform and would reduce the risk in developing COVID-19 [36], it is expected that self-restraint in obeying these protocols will diminish over time among students [24] due to the students’ lack of human interaction, connection to peers and social events [37, 38] during community quarantines.

How long will students strictly follow health policies in the campus to guarantee low COVID-19 transmission if strict compliance is not guaranteed throughout the school year? The logistic decay functions (5) are utilized to model decreasing personal protection and physical distancing over time, and we assume students adhere to health protocols at the start of the school year (*t* = 0) where personal protection *α*(0) is at most 50% while physical distancing *d*(0) is at most 3 meters, but have weakened at the point of inflection for some time *t*_0_. Fig. 10 shows COVID-19 incidence rates for all proposed models where health protocols are strictly implemented for *t*_0_ days. For the 4-10 cycle model, it is at least 50-63 days that health policies are to be strictly implemented; but for the rest of the models, at least 84-126 days of strict adherence to these protocols resulting to low disease transmission (*IR <* 10).

**Figure 10:**
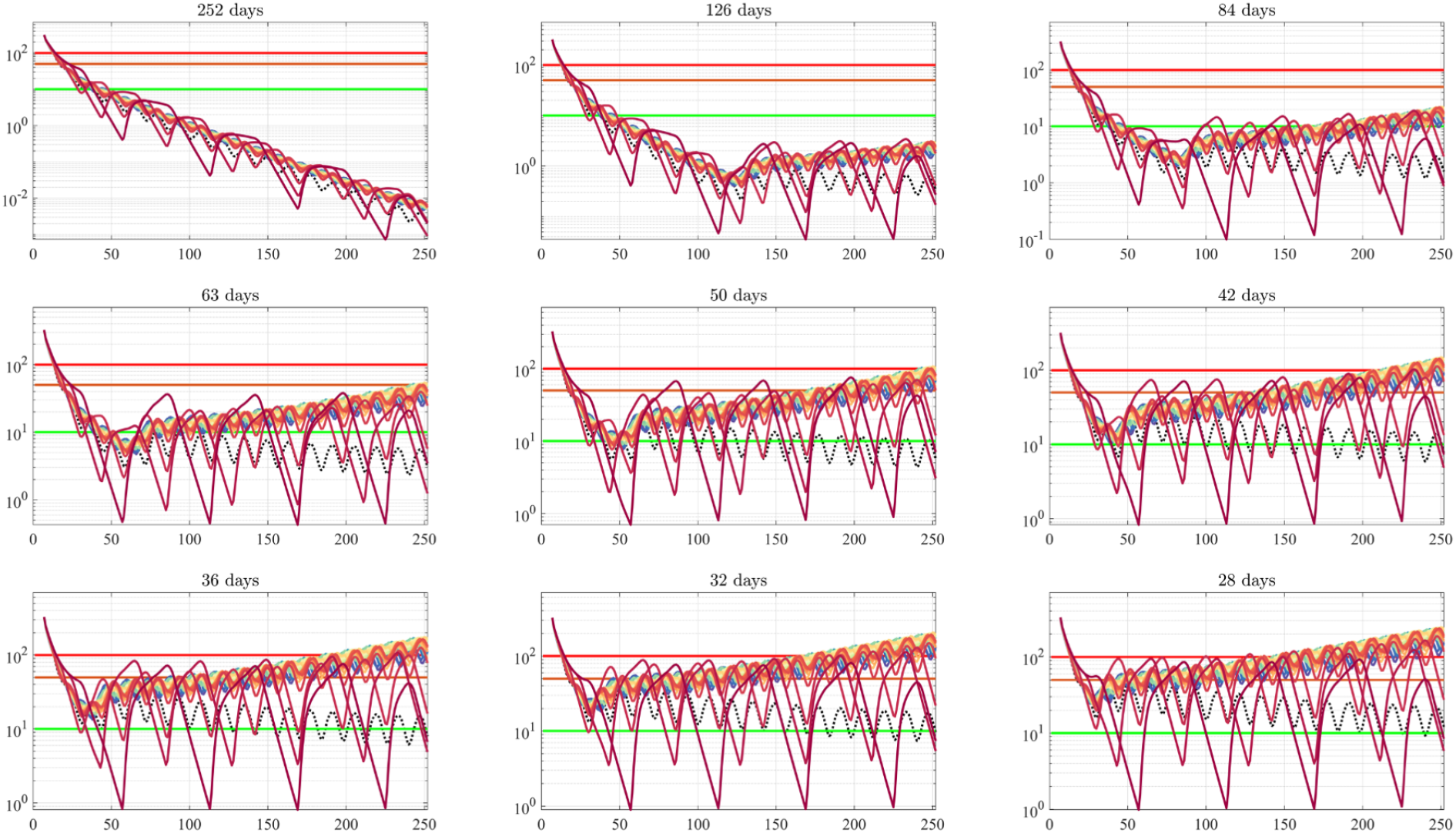
7-day IRs per 100 thousand for each cyclical schedule where health protocols are strictly implemented at most *t*_0_ days

**Figure 11:**
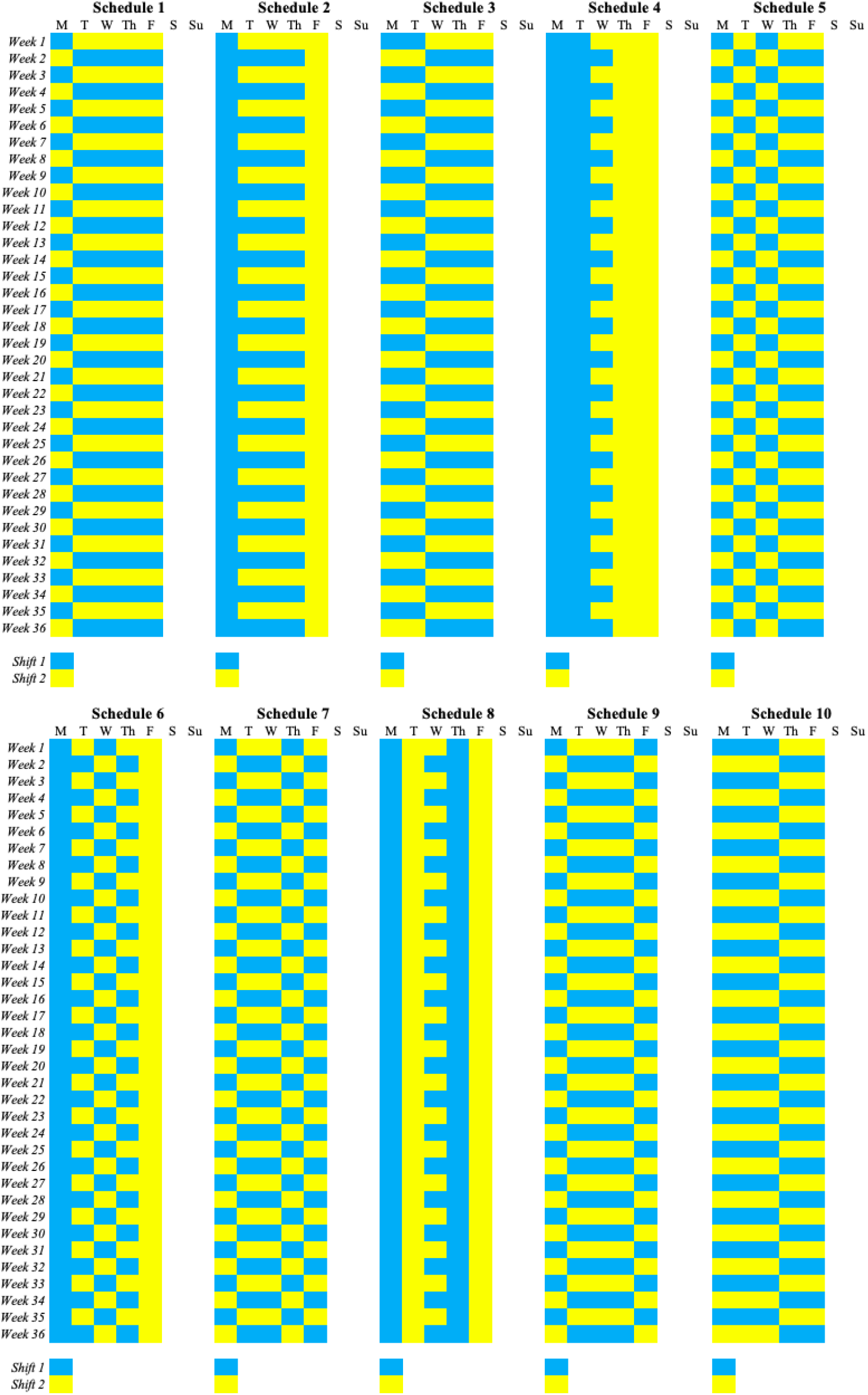
Cyclical Shifting Models: 1/4-day cycle (Schedules 1-2), 2/3-day cycle (Schedules 3-9), 3/2-day cycle (Schedule 10)

**Figure 12:**
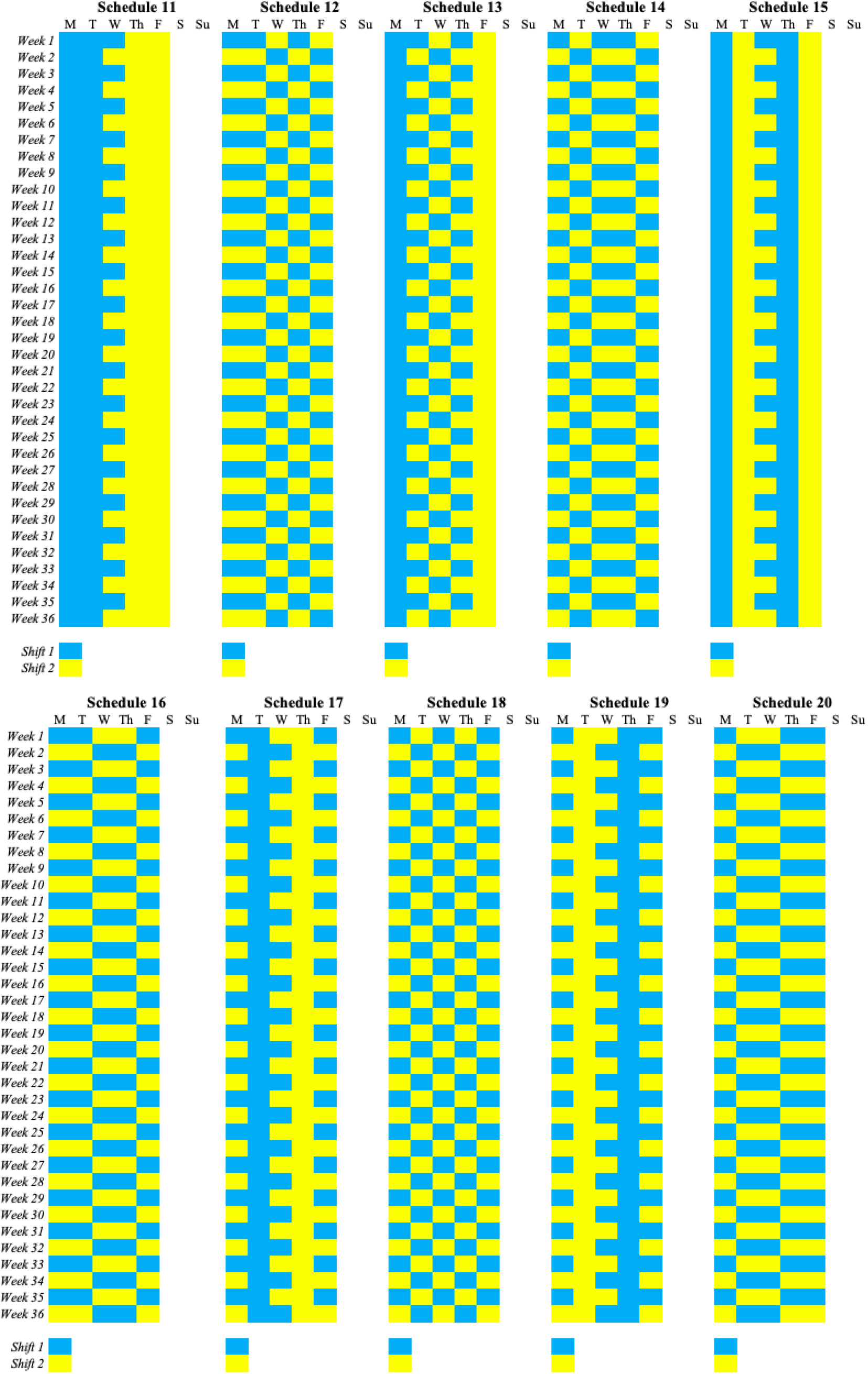
Cyclical Shifting Models: 3/2-day cycle (Schedules 11-20)

**Figure 13:**
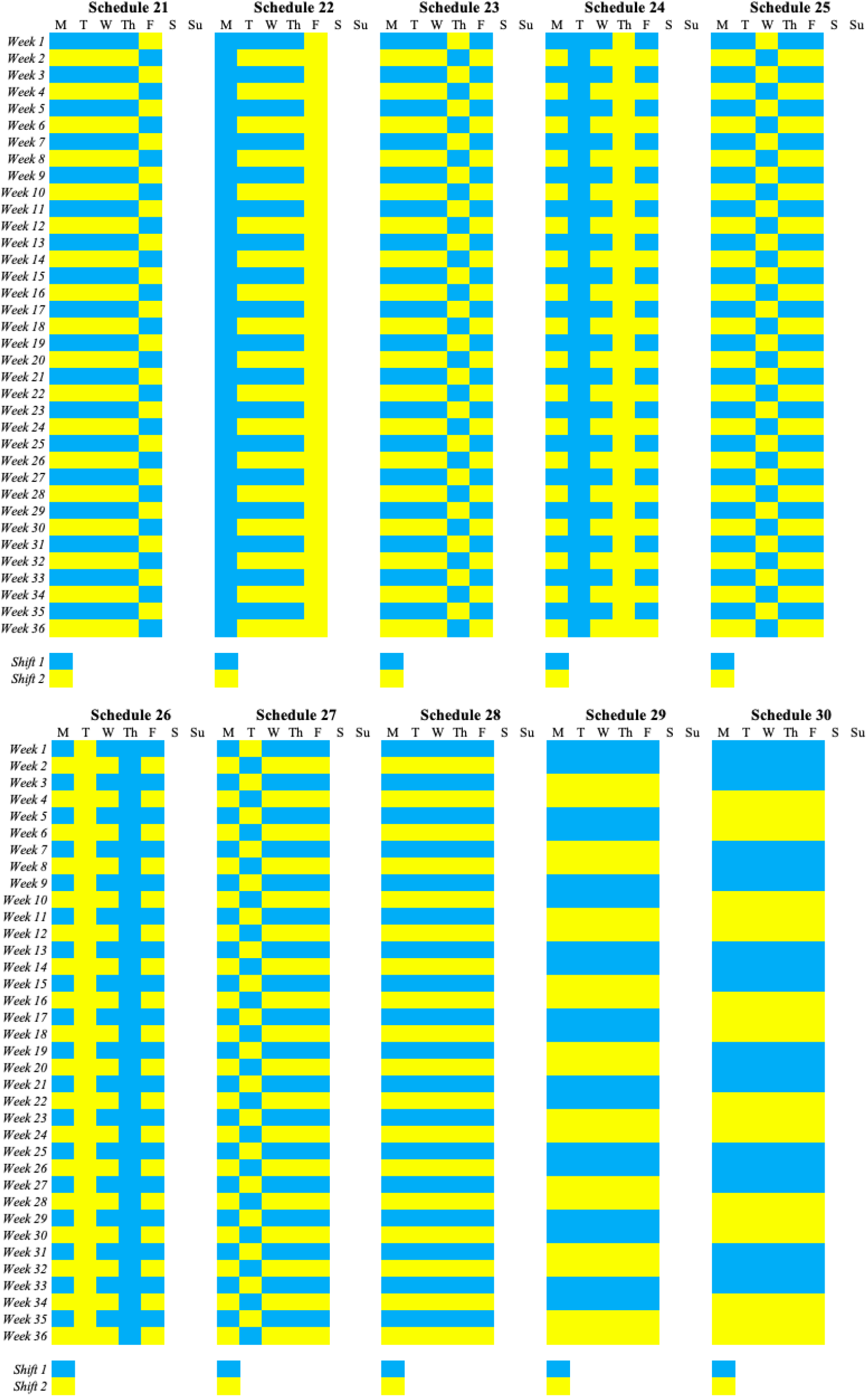
Cyclical Shifting Models: 4/1-day cycle (Schedules 21-27), 5-9 cycle model or 5/0-day cycle (Schedule 28), 2-week cycle (Schedule 29), 3-week cycle (Schedule 30)

**Figure 14:**
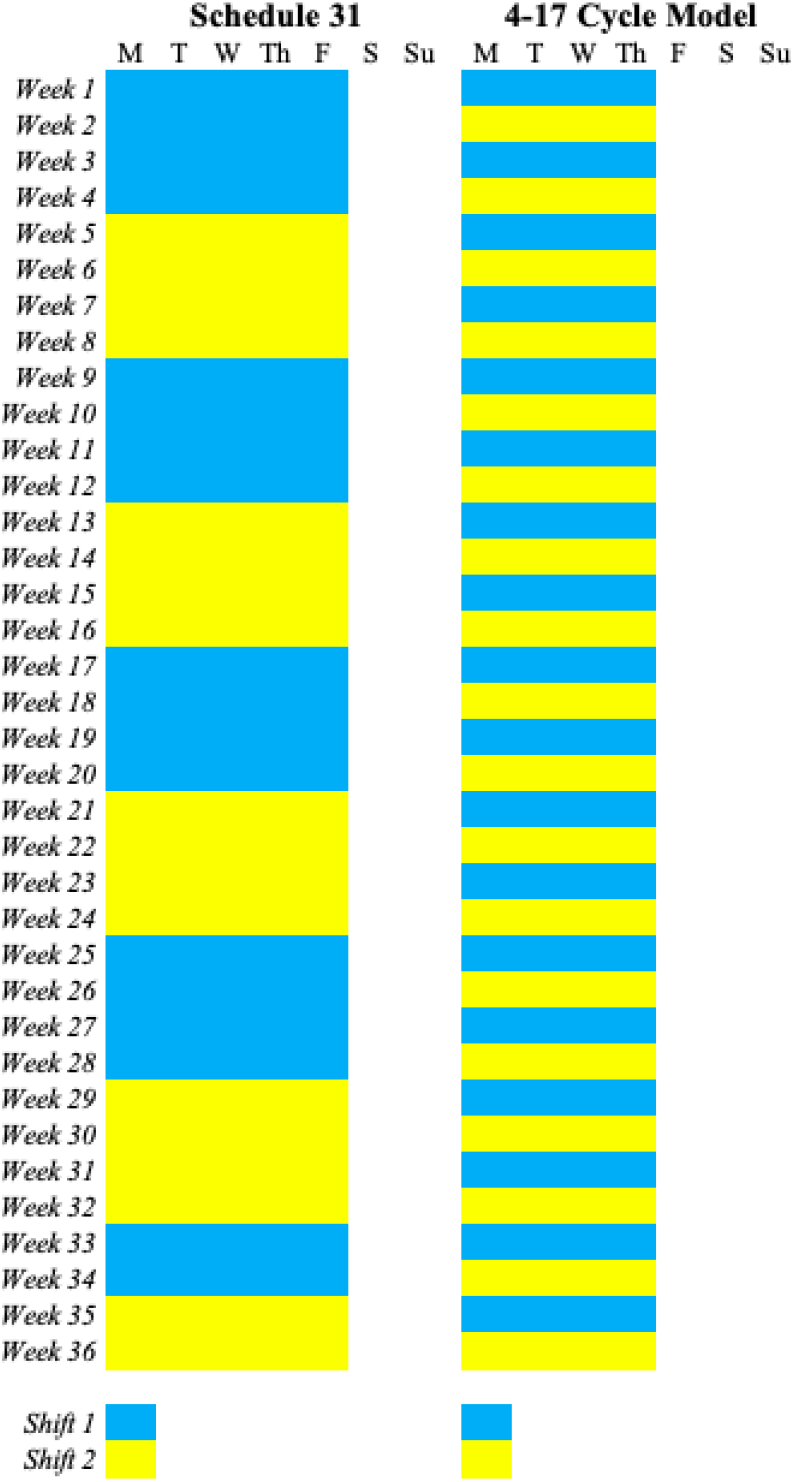
Cyclical Shifting Models: 4-week cycle (Schedule 31), 4-10 cycle model

## 4 Conclusion

The purpose of this simulation study was to investigate the effect of cyclical student scheduling models coupled with personal protection and physical distancing on the incidence of COVID-19 in a limited face-to-face scenario. Together with the implementation of health protocols such as mandating students to effectively wear protective gears such as face masks and the implementation of physical distancing, these combination of NPIs were the proposed basic health measures that will be implemented by colleges and universities in the Philippines when in-campus classes and activities are allowed [7].

Our simulation has shown that the recommended 4-10 cycle model in the joint CHEDDOH memorandum circular is an effective cyclical shifting schedule that schools can adapt; however, it is only limited to a 4-day face-to-face class every two weeks and about one-third of the competencies are taught in-campus. This paper also considered 2-shift schedules that would allow the teaching of about 50% of the competencies in face-to-face instruction. These schedules are classified as *m/n*-day cycle models (Schedules 1-28), 2-week cycle model (Schedule 29), 3-week cycle model (Schedule 30) and 4-week cycle model (Schedule 31).

Based on the simulations, the 3-week and 4-week cycle models can be adapted as cyclical shifting schedules if many students will less likely obey school policies on COVID-19 protection. However, if these policies are strictly implemented throughout the school year, any of the proposed schedules are effective in lowering disease transmission.

Moreover, this paper also investigated the effect of a declining students’ adherence to policies and answer the question on how long these policies to be sustained to guarantee low disease transmission in the event that students may not able to strictly follow the mandated masking and physical distancing. Findings point to at least half the school year of strict implementation would yield low transmission.

This paper has supported the joint CHED-DOH policies on limited face-to-face classes. This will likely help and guide school officials on the choice of a cyclical shifting model; in fact, the results of this work have been used by the authors as a contribution to the formulation of the COVID-19 academic plan of their institution for a resilient and safe re-entry of students in the campus.

## Data Availability

All data produced in the present work are contained in the manuscript.

https://www.mathworks.com/matlabcentral/fileexchange/115835-cyclical-student-shifting-models-in-the-time-of-covid-19

## Supplementary Materials

The ^∗^.*m* files containing the codes can be downloaded from A.J. Balsomo’s MATLAB File Exchange.

## Acknowledgment

The authors were supported by the University Research and Development Center, West Visayas State University. A.J. Balsomo acknowledges the Ateneo School of Medicine and Public Health, and Public Health England for hosting the workshop on *Mathematical Modeling for UHC in the Philippines*, supported by a Researcher Links Workhop grant ID RLWK11-10768 under the Newton-Agham partnership.

## Appendix

## Notes

### Competing Interest Statement

The authors have declared no competing interest.

### Funding Statement

This study was funded by the URDC of the West Visayas State University.

